# Validating and automating learning of cardiometabolic polygenic risk scores from direct-to-consumer genetic and phenotypic data: implications for scaling precision health research

**DOI:** 10.1101/2022.03.01.22271722

**Authors:** Arturo Lopez-Pineda, Manvi Vernekar, Sonia Moreno Grau, Agustin Rojas-Muñoz, Babak Moatamed, Ming Ta Michael Lee, Marco A. Nava-Aguilar, Gilberto Gonzalez-Arroyo, Kensuke Numakura, Yuta Matsuda, Alexander Ioannidis, Nicholas Katsanis, Tomohiro Takano, Carlos D. Bustamante

## Abstract

**Introduction:** A major challenge to enabling precision health at a global scale is the bias between those who enroll in state sponsored genomic research and those suffering from chronic disease. More than 30 million people have been genotyped by direct-to-consumer (DTC) companies such as 23andMe, Ancestry DNA, and MyHeritage, providing a potential mechanism for democratizing access to medical interventions and thus catalyzing improvements in patient outcomes as the cost of data acquisition drops. However, much of these data are sequestered in the initial provider network, without the ability for the scientific community to either access or validate. Here, we present a novel geno-pheno platform that integrates heterogeneous data sources and applies learnings to common chronic disease conditions including Type 2 diabetes (T2D) and hypertension.

**Methods:** We collected genotyped data from a novel DTC platform where participants upload their genotype data files, and were invited to answer general health questionnaires regarding cardiometabolic traits over a period of 6 months. Quality control, imputation and genome-wide association studies were performed on this dataset, and polygenic risk scores were built in a case-control setting using the BASIL algorithm.

**Results:** We collected data on N=4,550 (389 cases / 4,161 controls) who reported being affected or previously affected for T2D; and N=4,528 (1,027 cases / 3,501 controls) for hypertension. We identified 164 out of 272 variants showing identical effect direction to previously reported genome-significant findings in Europeans. Performance metric of the PRS models was AUC=0.68, which is comparable to previously published PRS models obtained with larger datasets including clinical biomarkers.

**Discussion:** DTC platforms have the potential of inverting research models of genome sequencing and phenotypic data acquisition. Quality control (QC) mechanisms proved to successfully enable traditional GWAS and PRS analyses. The direct participation of individuals has shown the potential to generate rich datasets enabling the creation of PRS cardiometabolic models. More importantly, federated learning of PRS from reuse of DTC data provides a mechanism for scaling precision health care delivery beyond the small number of countries who can afford to finance these efforts directly.

**Conclusions:** The genetics of T2D and hypertension have been studied extensively in controlled datasets, and various polygenic risk scores (PRS) have been developed. We developed predictive tools for both phenotypes trained with heterogeneous genotypic and phenotypic data generated outside of the clinical environment and show that our methods can recapitulate prior findings with fidelity. From these observations, we conclude that it is possible to leverage DTC genetic repositories to identify individuals at risk of debilitating diseases based on their unique genetic landscape so that informed, timely clinical interventions can be incorporated.

## 1. Background

Early diagnosis and prevention of chronic modern diseases, including type 2 diabetes (T2D) and hypertension, have the potential to make a significant impact in patient outcomes. However, the Centers for Disease Control (CDC) estimated that over 20% of T2D cases are undiagnosed [1] and that only 11% of the over 80 million U.S. residents that suffer from prediabetes have been diagnosed (CDC National Diabetes Statistics Report 2017). Early diagnosis could allow for better allocation of intervention strategies known to be effective at reducing the risk of disease progression. According to medical practitioners, insufficient screening is lacking mainly due to the fact that chronic diseases tend to progress slowly until they manifest clinically later in life. One of the main barriers to effectively identifying individuals at risk is the lack of predictive tools trained on heterogeneous datasets that are able to predict susceptibility using historical data available outside of clinical and research settings.

The World Health Organization (WHO) reports a sustained increase in diabetes mellitus, with projections increasing to 3% of the world population by 2030, becoming the seventh leading cause of death globally [1]. A sedentary lifestyle and a diet pattern with high intake of foods rich in hydrogenated fat, refined grains, and red meat have contributed to the increase in overweight and obesity and led to the increased incidence of T2D [2]. An important challenge to this health crisis is to decrease mortality, especially at younger ages, and in low and low-middle countries [3], with more than 400 million people affected globally [4].

The overlap between T2D and hypertension is common among the population [5]. Hypertension alone affects more than 1.28 billion people worldwide [6]. T2D can lead to complications which can be exacerbated when the patient also presents hypertension, for example in the progression of diabetic nephropathy [7]. Both T2D and hypertension are risk factors associated with stroke and other serious and life-threatening events [8]. In fact, during the recent COVID-19 pandemic, outcomes of patients seem to be negatively affected by the presence of T2D, hypertension and obesity [9].

Genetics of T2D has been extensively studied [10-16], with over 400 genetic variants found to be associated with the diseases [17]. In addition to individual studies focusing on defined ethnic groups like Hispanics [18, 19], there have been consortium efforts to investigate the genetic architecture of complex traits in diverse populations. Some of these consortia include a) the Population Architecture using Genomics and Epidemiology (PAGE) consortium [20]; b) the rich data offered by the UK Biobank allowing associations between complex traits, genetics, and lifestyle [21]; c) the Trans-Omics for Precision Medicine (TOPMed) Consortium [22] which improves imputation quality and detection of rare variant associations; and more recently d) the Meta Analysis Biobank Initiative [23], a collaborative network of biobanks across the world representing millions of consented individuals.

Direct to consumer platforms are novel sources of information that have expanded quickly during the past decade. The earliest example in 2010 was the use of web-based self-reported questionnaires with complementary genetic testing, leading to the creation of a research database [24] which has allowed for novel polygenic risk scores in complex traits [25] and subsequent FDA submissions of novel diagnostics [26]. The clinicogenomic database developed by a consortium led by a large pharmaceutical company, alongside an electronic health record company focused on oncological practices, and a direct to consumer (DTC) genetic testing company, putting together a comprehensive database [27], allowing sophisticated analysis of including the selection of novel biomarkers [28], drug effectiveness studies [29], or automatic eligibility criteria selection [30].

The rapid development of polygenic risk scores (PRS) in recent years stresses the importance of accurately assessing the ancestry makeup of participants in biomedical studies to avoid potential selection biases [31]. PRS represents a measure of an individual’s overall genetic liability to a trait or disease [32]. The European Bioinformatics Institute (EBI) has launched a PRS catalog database, allowing for reproducibility and standardization of reporting of PRS models [33]. As of Feb 16, 2022, the catalog includes 35 models related to the T2D from 15 peer reviewed publications [34-46], and six models related to hypertension across 3 publications [39, 44, 46]. However, these models mostly include single ancestry participants (typically European) which may not generalize across other ancestral groups.

Even though the need for ancestry-focused research has been highlighted by many [47, 48], the lack of diversity resulted in systemic biases that threaten to widen existing health disparities among minority and majority populations in most developed countries. The overrepresentation of European individuals in genetic studies represents a major issue, hampering the translation of PRS across populations. In this study, we hypothesize that PRS models can be improved by defining the genetic ancestry of participants. Embedding genetic ancestry as a covariate, or scaling PRS scores as part of post-processing step, would result in more accurate models than traditional filtering to only European-based PRS models.

Furthermore, the validation of a DTC framework for validating and extending PRS provides a cost-effective means of enrolling understudied populations in complex disease genetics as 23andMe has done with their “Roots into the Future” effort. We aim to build on such successes by powering a public-private partnership that is collecting DTC data to be included in the Biobank Meta-analysis Network. The notion that DTC provided a “straight to mobile instead of landlines” opportunity is important to validate as most low and middle income countries are considering how to harness advances in genomics for the study of their own populations while building state-sponsored capacity is a fundamental challenge to many efforts.

## 2. Methods

In this case-control retrospective observational study, adult participants from an international genetic platform were invited to self-report their health status and metabolic traits. Their genetic information was also previously uploaded in the same platform, which allowed us to explore their genetic susceptibility and to build polygenic risk scores (PRS) regarding these traits. Finally, we calculated ancestry estimation using Neural ADMIXTURE for all individuals. We were interested in evaluating ancestry-aware polygenic scores for type 2 diabetes and hypertension.

### 2.1 Cohort and Eligibility Criteria

We used the research database where participants were drawn from Genomelink (genomelink.io) users, which offers a DTC genetic traits platform with more than 500,000 users globally. After uploading their genetic information, generated in other DTC platforms, users can be informed on their susceptibility to an extensive set of genetic traits. All participants created an account, and agreed to a Consent on the use of their data and Legal Agreement. Upon signing up, participants were invited to undertake a health online survey. Participants were redirected to the survey once they gave online consent to be a part of the research. The online consent is in compliance with the institutional review board (IRB) at WCG IRB (https://www.wcgirb.com/) under IRB tracking ID 20201332.

The online survey included questions about general conditions like diabetes, blood pressure, lipid profile, and medication intake. It also included COVID-19, influenza and common cold-related questions along with age, sex, weight, height, and pandemic behavior. Data were collected over a period of six months, from May 01, 2021 to October 06, 2021. Supplementary Material A shows the online questionnaire.

Only the initial answers of each participant were included in the study, if genotype information was available, and if they answered the age and sex questions. Case-control groups were created following participants’ answers to the general condition question: T2D and hypertension. Additionally, for T2D, participants were included if they reported to have high levels of sugar in their blood work or if they were taking antidiabetic medication. Participants were defined as controls if they did not report managing health conditions listed in the questionnaire survey, or if they were not managing any health condition. Also, for the T2D cohort specifically, participants who reported having normal sugar levels were included as controls. Participants with missing values were excluded.

### 2.2 Genotype data: quality control, imputation and GWAS

This study includes seven independent genotyping arrays, comprising a total of 12,424 unrelated individuals. Genotype-level data for each array were processed by applying identical quality control and imputation procedures. Briefly, variants with a call rate of <95% and palindromic markers (A/T, G/C, MAF > 0.4) were excluded. We performed an exact test for Hardy– Weinberg equilibrium for individuals of the largest ancestral group (*p*□< □1□×□10^−12^, globally). Individual quality control (QC) includes genotype call rates >97%, matching between gender identification and chromosomal sex, and no excess ancestry-adjusted heterozygosity. Samples genetically related to other individuals in the cohort and duplicates were detected and removed, by applying the King algorithm (--make-king, king estimate > 0.177; PLINK 2). Principal component analysis was performed to identify global ancestry per individual using 1000 genomes as reference population with PLINK 2 [49]. Further information about the number of markers per genotyping array pre- and post-QC is available on Supplementary Material B.

Imputation was carried out using 1000 genomes as a reference panel with Beagle [50]. Next, we generated a merged data set combining imputed genotypes (MAF >0.01; imputation quality *R*^2^□>□0.30) from available data sets. Imputed makers with call rate >0.95 in the merged data were selected for downstream analysis.

The GWAS was performed for T2D and hypertension phenotypes (N = 4,550; N = 4,528; respectively) using an additive genetic model with PLINK 2 (--glm). We include the top ten principal components (PC)s, age, sex, and the genotyping array as covariables in the model. Results were depicted using the qqman package in R.

### 2.3 PRS analysis

The Batch Screening Iterative LASSO (BASIL) algorithm [51] is a meta-algorithm (algorithms that learn from the output of other algorithms), which employs a Lasso algorithm [52] and enhances this output with another layer for faster variable selection in ultra high dimensional problems. Similar to the Lasso algorithm, the purpose of BASIL is to find a parameter vector L whose components are the coefficients for the independent variable of the linear regression that approximates the solution of the problem.

BASIL solves the Lasso solution path in an iterative fashion, starting with a sequence of candidate parameters. From these candidate solutions, each iteration discards the ones that do not meet the requirements to be a suitable solution. The set of variables who make it into the final set for a viable solution are those who were also screened satisfying a desired threshold requirement, while the others are discarded (i. e. those solutions in which the coefficients in their positions inside the □ parameter are meant to be 0). This process is repeated until the optimum parameter □0 is found, which is the one that minimizes □ (□0). The BASIL algorithm guarantees to find the exact solution and not only an approximation, via the Karush-Kuhn-Tucker condition (the first derivative necessary conditions for a solution to be optimal) [53] which is verified along each iteration. This condition is necessary and sufficient to prove it.

### 2.4 Genetic ancestry

To address the confounding factor of population stratification in PRS estimations, various approaches have been taken [54, 55]. Here, we follow the convention of using the first 10 principal components of the PCA to the adjustment of the GWA study. For the correction of PRS models, we make use of estimates of global ancestry. For this purpose, we use Neural ADMIXTURE [56], a faster adaptation of the ADMIXTURE algorithm [57] with similar (or better) clustering results. Utilizing the Python implementation of Neural ADMIXTURE, we use data from the 1000 Genomes Project Consortium [58] for training a model in the supervised mode of Neural ADMIXTURE with the default parameters. We utilized the results of global ancestry inference as a covariate in the training of our PRS models.

### 2.5 Statistical analysis

We first used descriptive statistics to provide baseline characteristics of the study participants. We built two PRS models using a mixed model regression. In each PRS the predictor variable was taken from the online questionnaire, as mentioned in the inclusion criteria. A binary prediction for each phenotype was calculated, while accounting for sex, age and the first ten genetic principal components (PCs). These PCs account for residual population micro-stratification as fixed effects. Genetic relatedness matrix was built using PLINK 2 (kinship estimation), to account for the relatedness among individuals as a random effect.

The predictive ability of these PRS models was evaluated using the area under the curve (AUC) receiver operating characteristic (ROC) curves using the pROC package in R [59]. To make comparisons between AUC curves from each model, we used the nonparametric method developed by DeLong et al. [60], which is a commonly used method.

## 3. Results

### 3.1. Genome-wide association results for Diabetes type 2 and Hypertension

We combined genome-wide association data for: 1) 389 T2D cases and 4,161 controls (N = 4,550); and 2) 1,027 hypertension cases and 3,501 controls of European ancestry (N = 4,528). We tested ∼8 million variants for T2D and hypertension passing quality control and imputation filters (MAF>0.01, *R2*>0.3). Both results showed low inflation of test statistics (λGC□ □= □ □1.01; λGC□ □= □ □1.01) (Figure 1).

**Figure 1.**
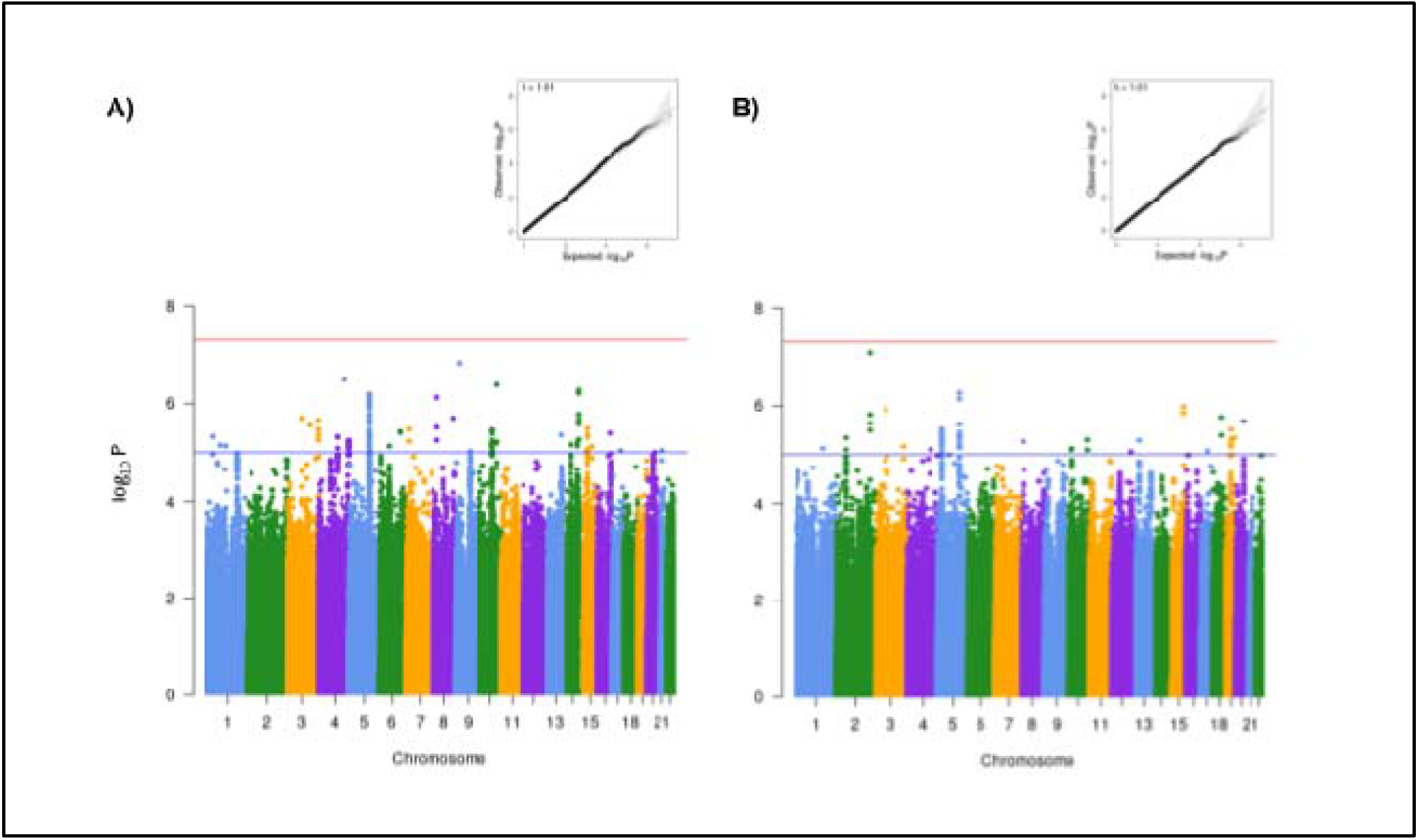
Genome-wide association results. A) type 2 diabetes, and B) hypertension. Top boxes show Q-Q plots, while bottom figures show Manhattan plots with two levels of significance of p < 5 × 10^−8^ (red line), and p < 1 × 10^−6^ (blue line)

Nineteen T2D variants displayed significant evidence of replication (p < 0.05) in this dataset. Among them, we identified variants closely associated with genes which have been previously linked to type 2 diabetes susceptibility (e.g. *CDKAL1, KCNQ1*), as well as variants in the *FTO* locus linked previously with both BMI and T2D. Overall, we identified 164 out of 272 variants showing identical effect direction to previously reported genome-significant findings in Europeans (Supplementary Material C) [36]. For the hypertension dataset, we replicated ten hypertension genetic markers, and identified 230 out of 365 variants having identical effect direction [61] (Supplementary Material C; Figure 1).

We validated our GWAS using independent GWAS meta-analysis datasets from Mahajan et al. 2018 (74,124 T2D cases, 824,006 controls) [36] and Evangelou et al. 2018 (757,201 individuals) [61]. We compared the p-values and the effect sizes for the variants assessed in both our studies that had identical chromosomal coordinates and alleles with the independent GWAS. The direction of the effect sizes (estimated as OR) were set to match the effect alleles in each study. We observed that the effect sizes of the genome-wide significant variants in the independent GWAS [36, 61] were concordant in directionality in both our T2D and hypertension GWAS (effect sizes had the same direction across both studies, Supplementary Material C).

Our observations highlight how carefully curated DTC repositories with ever increasing sample sizes and variant diversity can replicate previous findings and hold the potential of delivering enhanced discovery and single-variant resolution of causal T2D and hypertension risk and protection alleles. Additionally, our findings confirm the potential impact of DTC resources on mechanistic insights and clinical translation efforts.

### 3.2 Estimation of cardiometabolic PRS models using SNPnet

PRS models were built for each phenotype using the BASIL algorithm [51]. The predictor variable was binary (presence or absence of diabetes or hypertension) as reported by participants. After 10-fold cross validation, the models reported a predictive performance (measured using AUC) of 0.68 for both diabetes and hypertension. Similarly, the models built when filtering only for participants of European ancestry reported a predictive performance of 0.69 and 0.66 respectively. We compared the performance of these models using DeLong’s method [60] with no statistical significance (See Supplementary Material D for additional metrics). The imbalance ratio of both cohorts was an important factor that impacted the accuracy of these models. Additionally, the majority proportion of participants of mostly European ancestry also explains the small differences in performance between both types of models. Figure 2 shows the comparison between AUC curves in both phenotypes.

**Figure 2.**
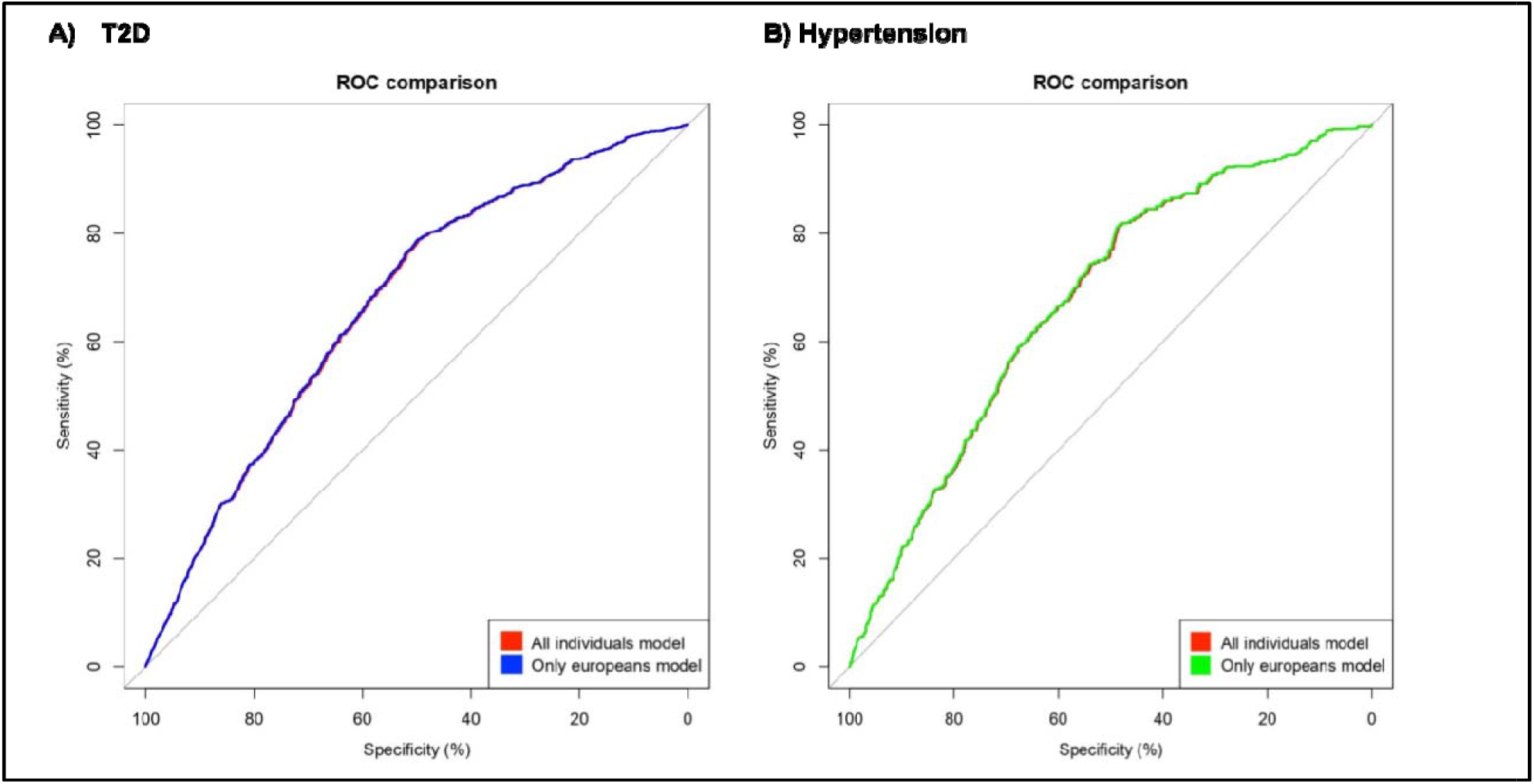
Area under the curve (AUC). Comparison of receiver operating characteristic (ROC) between two models Full model and European only model. Results for A) type 2 diabetes (T2D) were 0.68 and 0.69 respectively; while for B) hypertension were 0.68 and 0.66 respectively. After applying the DeLong [60] method of ROC comparison the models were not significantly different.

The European Bioinformatics Institute (EBI) developed the PGS Catalog [32], which is an open resource of published polygenic scores (including variants, alleles and weights). We investigated those published PRS for T2D and hypertension. For those with reported AUC, we also obtained the number of variants, number of individuals whose data was used to train the model under various ancestry groups. See Supplementary Material E for more information on the PGS Catalog reported scores.

Our models are comparable to previously published PRS models. Figure 3 shows the comparison in reported AUC for those models in the EBI PGS catalog including our own models. The average AUC between these models was 0.70. The small number of variables used by our models (125 for T2D and 666 for hypertension), makes them comparable to those reported by Tanigawa et al. [44] who also used the BASIL algorithm. Likewise, the number of individual whose data was used to train the models is modest in comparison to large academic and clinical databases. Nevertheless, the predictive performance does not seem to be overtly affected by the number of individuals in the study or the number of included variants highlighting that genetic array data from DTC repositories carry immense promise for the development of PRS tool aimed at improving early detection and prevention of T2D and hypertension.

**Figure 3.**
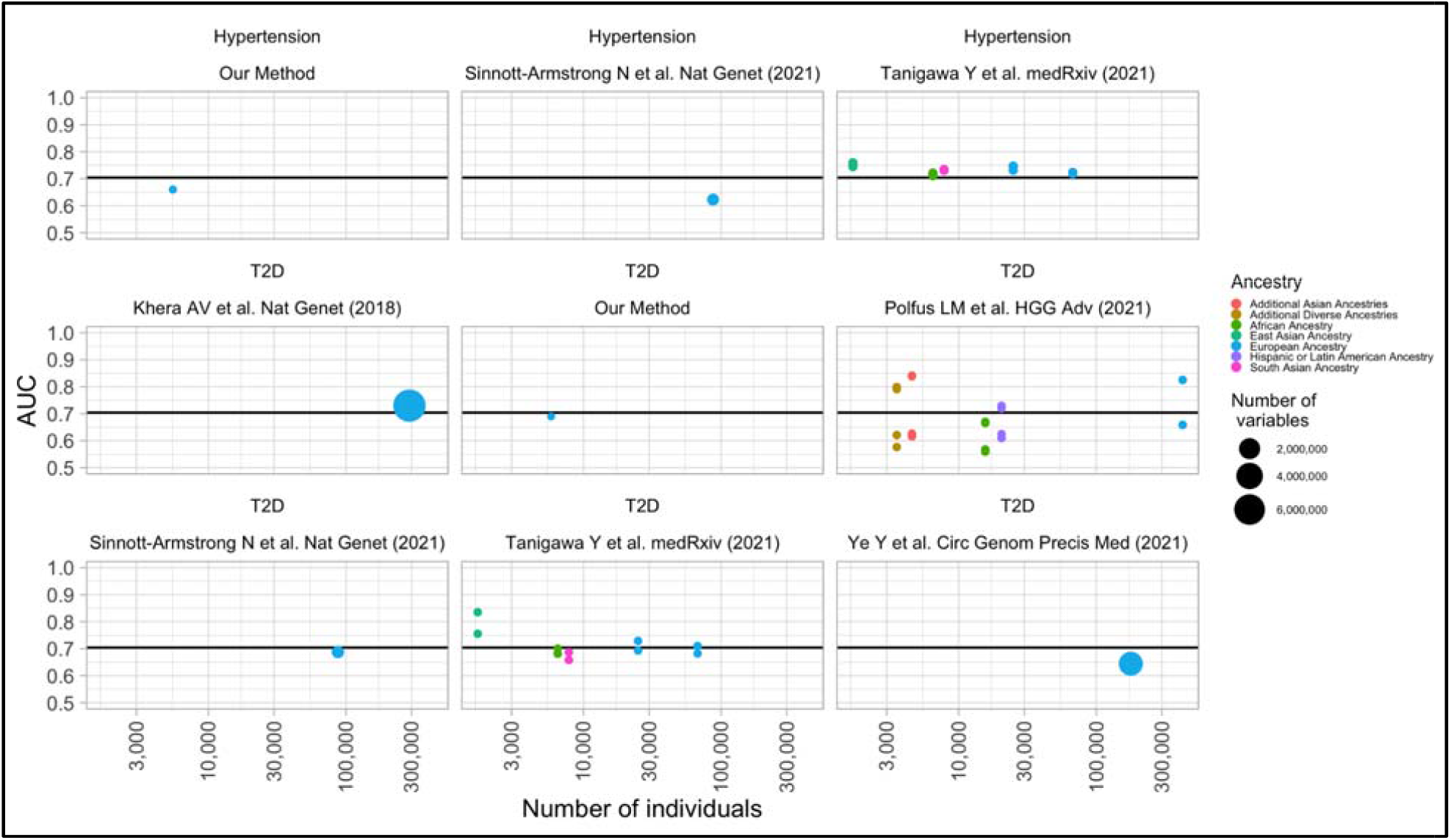
Comparison of PRS published in the EBI PGS Catalog for T2D and Hypertension. The color of the bubble represents the population ancestry that was included to build the PRS model. The size of the bubble represents the number of variables (variants) that ended up in the model after training. The x-axis shows the number of individuals used to train the model. The y-axis shows the AUC results as reported in the EBI PGS Catalog. The horizontal line shows the average AUC across all models.

## 4. Discussion

In this study we generated PRS for T2D and hypertension from a heterogeneous dataset housing a combination of genetic data and self-reported information from a DTC genetics company. Despite a relatively modest predictive ability our PRS models are able to identify subsets of users at substantially increased risk of presenting T2D or hypertension. This finding is remarkable because it suggests that the ever increasing availability of genetic data from DTC providers, most of it not annotated for traits of clinical relevance, can be leveraged to generate predictive tools able to improve diagnosis and prevention of diseases with genetic determinants.

Our study tested the possibility of inverting the model regarding genetic and phenotypic data acquisition in a research study. Individuals participating in our study shared their genetic array information from other DTC providers, and were invited to take an online survey regarding their general health condition. We found no difference in predictive performance between our trained models that included respondents from all inferred ancestries and those models with respondents from European heritage, due to the fact that 86% of our database was of European origin. The genetics of our PRS models for T2D and hypertension are supported by our ability to replicate known variants from publicly available independent GWAS studies.

Multiple array types were available in our database, and imputation across platforms (up-imputing) was necessary to harmonize these diverse datasets. The fact that individuals can self-report their genomic information could potentially corrupt the file being uploaded into our platform. However, applying appropriate quality control (QC) principles proved to successfully enable traditional GWAS and PRS analyses.

DTC platforms can offer a wide range of information about personal wellness, ancestry, physical characteristics, and traits. Advances in genomic research have led the DTC genomics industry to flourish and make accurate yet easy to interpret genomic results. Strict privacy policies of many companies disallow them to share customers’ data without their consent. These platforms can serve as informative repositories giving actionable insights that aid traditional clinical approaches. The approach of subject recruitment for various complex phenotypes via online surveys is opening up multiple avenues to complement conventional research and clinical strategies. DTC platforms also provide convenience along with a wider reach to recruit participants from various locations. They surpass barriers of single-point data collection centers to language restrictions thus allowing the aggregation of data from places with different ancestries and demographics. Democratizing the access to these genetic platforms and prediction tools will likely boost progress in precision medicine. In the future, we plan to investigate how federated learning approaches can further improve the possibility to increase the power of studies in DTC genomic analysis, but also how meta-analysis can be done in combination with academic and clinical datasets (including those from large consortiums).

We have shown that our DTC platform and research strategy has the potential to replicate previously reported results with a very fast turnaround time. The participation of individual customers in our platform allowed the generation of a rich dataset that enabled the creation of PRS cardiometabolic models. The comparable predictive performance of our models also is a great indication of how we can quickly contribute more PRS models to the larger scientific community.

T2D and hypertension are multifactorial diseases that are impacted by genetic and environmental determinants, including lifestyle factors like nutrition and exercise habits. Therefore, providing personalized information about T2D and hypertension predisposition is poised to improve early diagnosis and prevention bringing precision medicine at scale for all.

## Data Availability

The data that supports the findings of this study is available for qualified researchers at non-profit institutions upon entering into an agreement with Genomelink. All information will be shared subject to the above criterion upon request via.

## Acknowledgements

We are grateful to all our participants for being a part of this study.

## Competing interests

ARM, SMG, MTML, ALP, CDB, AI, MNA and NK are employees of or consultants to Galatea Bio. MV, KN, YM, and TT are employees of Genomelink. CDB, IA, and NK are shareholders of Galatea Bio stock. CDB, KN, YM, and TT are shareholders of Genomelink stock. The remaining authors declare that there is no conflict of interest regarding the publication of this article.

## Contributions

ALP, MV, SMG, and ARM designed the study. SMG, MNA, GGA, ARM, and performed analysis of data. BM, MTML, KN, YM, AI, NK, TT, and CDB provided interpretation of the results. ALP, ARM, SMG, and CDB drafted the manuscript, and all authors contributed critically, read, revised and approved the final version.

## Funding

This research is based on results obtained from a project, JPNP19001, subsidized by the New Energy and Industrial Technology Development Organization (NEDO)

## Ethics approval

This study was approved by the institutional review board (IRB) at WCG IRB (https://www.wcgirb.com/) under IRB tracking number protocol number 20201332.

