## Supplementary Material B for "Validating and automating learning of cardiometabolic polygenic risk scores from direct-to-consumer genetic and phenotypic data: implications for scaling precision health research"

Number of variants per genotyping platform pre- and pos-QC, and final genotyping call rate.

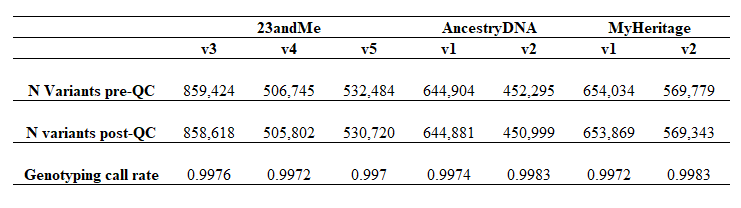
