## Supplementary Material D for "Validating and automating learning of cardiometabolic polygenic risk scores from direct-to-consumer genetic and phenotypic data: implications for scaling precision health research"

**Table D.1**. PRS models built with all participants (not filtered by ancestry) after 10-fold cross validation

| **Trait** | **Predictive performance (AUC)** | | | | **n** | **case_n** | **control_n** |
| --- | --- | --- | --- | --- | --- | --- | --- |
|  | **Geno** | **Covar** | **Full** | **Delta** |  |  |  |
| T2D | 0.56825 | 0.69478 | 0.68597 | -0.0088 | 5,818 | 474 | 5,344 |
| Hypertension | 0.56197 | 0.65216 | 0.68175 | 0.02958 | 5,526 | 1,204 | 4,322 |

**Table D.2**. PRS models built with participants of European ancestry after 10-fold cross validation

| **Trait** | **Predictive performance (AUC)** | | | | **n** | **case_n** | **control_n** |
| --- | --- | --- | --- | --- | --- | --- | --- |
|  | **Geno** | **Covar** | **Full** | **Delta** |  |  |  |
| T2D | 0.57822 | 0.69703 | 0.68912 | -0.0079 | 4,535 | 387 | 4,148 |
| Hypertension | 0.53188 | 0.655 | 0.65854 | 0.00354 | 4,513 | 1,023 | 3,490 |

**Geno**: the predictive performance of the predictive model with genotype features alone.

**Covar**: the predictive performence of the predictive model with covariates (age, sex) alone.

**Full**: the predictive performance of the full model that has both genotypes and covariates alone.

**Delta**: the incremental predictive performance of the full model against the covariate-only model, i.e. the difference between the values in "Full" and "Covar" columns.

**n**: the number of individuals

**case_n**: (binary traits only) the number of cases

**control_n**: (binary traits only) the number of controls
