## Supplementary Material F for "Validating and automating learning of cardiometabolic polygenic risk scores from direct-to-consumer genetic and phenotypic data: implications for scaling precision health research"

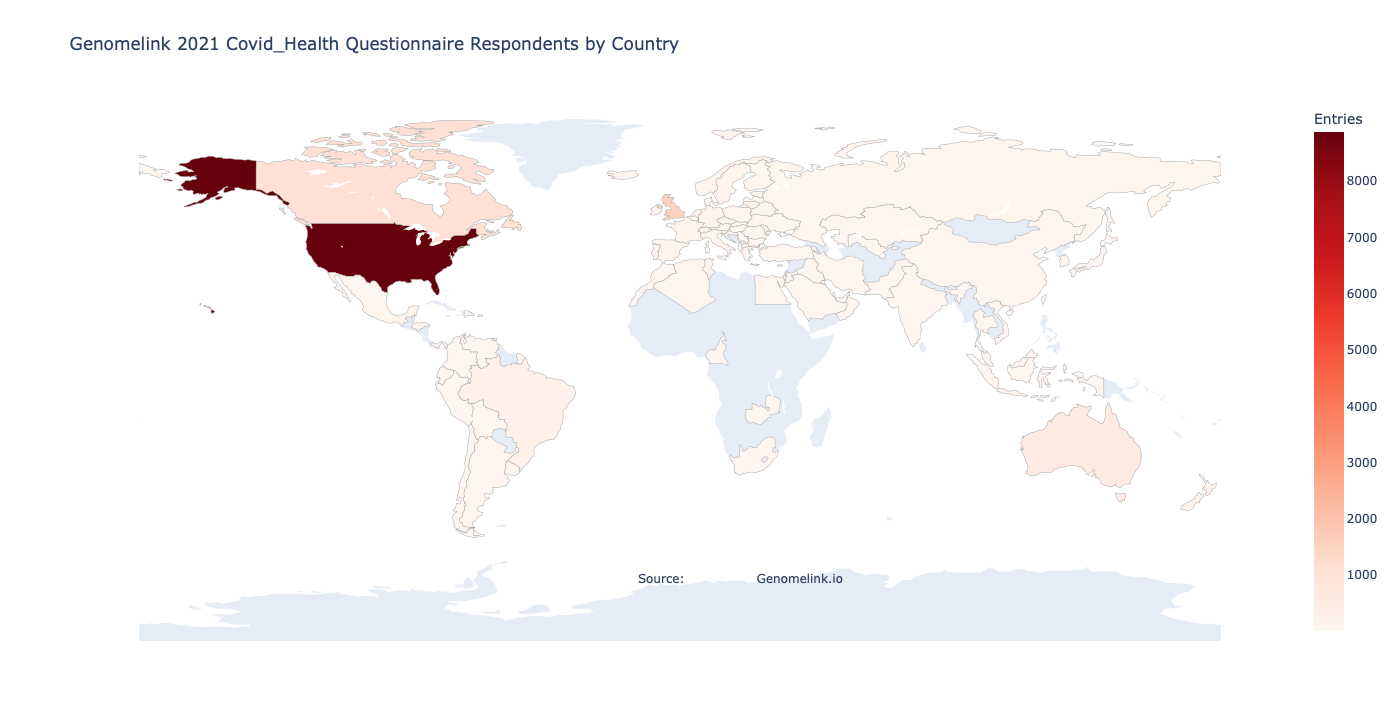

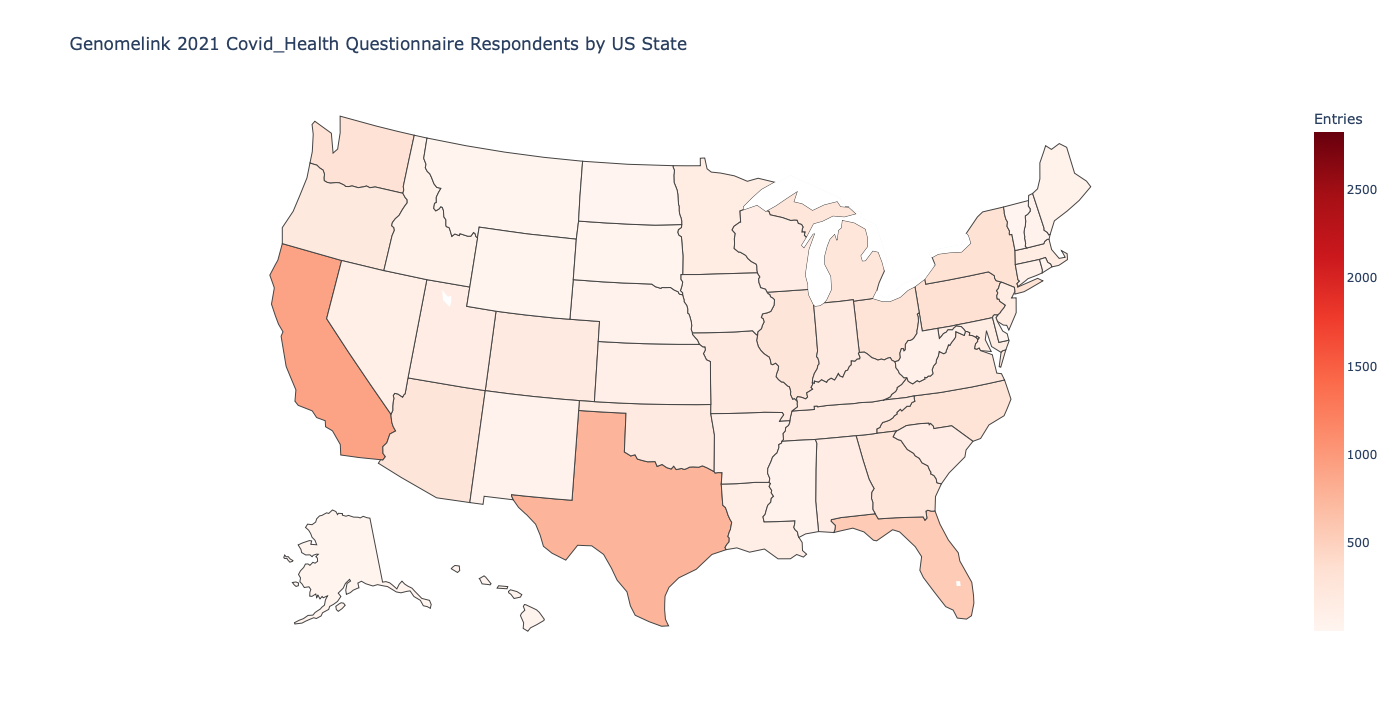

Q02021-10-08: 15391 respondents / 12424 unique

**Fig F.1. Distribution of respondents by geographic location.** Of the 15,391 entries of the questionnaire 12,424 correspond to unique respondents. Most respondents were located in the US (63.19%), followed by GB (11.17%) and CA (7.35%), with other participants responding from other locations on earth (18.27%). Most respondents were of European ancestry (86%). Participants were 61% female with 45% falling in the 45-64 age category.

|  | **Overall N (%)** |
| --- | --- |
| **N** | 12,424 (100%) |
| **Age (%)**  17 or younger  18-24  25-44  45-64  65 or older  Missing | 68 (0.6%)  636 (5.4%)  4,650 (39.4%)  4,778 (40.4%)  1,685 (14.3%)  607 (4.8%) |
| **Sex (%)**  Female  Male  Prefer not to say  Missing | 8,001 (71.7%)  3,072 (27.5%)  83 (0.7%)  1,268 (10.2%) |
| **Comorbidities**  Allergies  Not managing any health conditions at all  High blood pressure  Not managing any health conditions listed here  Arthritis  Asthma  Diabetes (Type 2)  Immune system suppression  Others  Missing | 2,579 (20.8%)  2,217 (17.8%)  2,015 (16.2%)  1,901 (15.3%)  1,409 (11.3%)  1,277 (10.3%)  786 (6.3%)  305 (2.5%)  1,456 (11.7%)  3,749 (30.2%) |
